# Relationships between wellbeing polygenic scores, brain structure, and psychopathology in children

**DOI:** 10.1101/2024.12.17.24319137

**Authors:** Christian K. Tamnes, Andreas Dahl, Dennis van der Meer, Ingrid Agartz, Dag Alnaes, Ole A. Andreassen, Kathryn L. Mills, Linn B. Norbom, Geneviève Richard, Ronald E. Dahl, Espen Røysamb, Lars T. Westlye, Lia Ferschmann

**Affiliations:** PROMENTA Research Center, Department of Psychology, University of Oslo, Oslo, Norway; Division of Mental Health and Substance Abuse, Diakonhjemmet Hospital, Oslo, Norway; Department of Psychology, University of Oslo, Oslo, Norway; Section for Precision Psychiatry, Division of Mental Health and Addiction, Oslo University Hospital & Institute of Clinical Medicine, University of Oslo, Oslo, Norway; School of Mental Health and Neuroscience, Faculty of Health, Medicine and Life Sciences, Maastricht University, Maastricht, The Netherlands; Institute of Clinical Medicine, University of Oslo, Oslo, Norway; Centre for Psychiatry Research, Department of Clinical Neuroscience, Karolinska Institutet & Stockholm Health Care Services, Stockholm Region, Stockholm, Sweden; K.G Jebsen Center for Neurodevelopmental Disorders, University of Oslo, Oslo, Norway; Department of Psychology, University of Oregon, USA; Institute of Basic Medical Sciences, Division of Anatomy, University of Oslo, Oslo, Norway; Institute of Human Development, University of California Berkeley, Berkeley, CA, United States; Department of Child Health and Development, Norwegian Institute of Public Health, Oslo, Norway; Norwegian Centre for Violence and Traumatic Stress Studies, Norway, Oslo, Norway

**Keywords:** ABCD Study, brain structure, children, intracranial volume, MRI, polygenic scores, psychopathology, wellbeing

## Abstract

Wellbeing is shaped by an interplay of genetic and environmental factors and is associated with both health and functioning. It remains unclear whether genetic influences on wellbeing are linked to brain structure and, in turn, early-life psychopathology. Here, we investigated associations between wellbeing polygenic scores (PGS), magnetic resonance imaging (MRI)-derived measures of brain structure, and parent-reported measures of child psychopathology in a large cross-sectional sample of children from the Adolescent Brain Cognitive Development (ABCD) Study (n = 8844; 8.9-11.0 years old). Preregistered analyses revealed no significant associations between wellbeing PGS and regional cortical thickness or subcortical volumes and only a small negative association with surface area of the parahippocampal cortex (β = −0.03, p=.002), while an exploratory analysis identified a small positive association with intracranial volume (ICV; β = 0.04, p<.001). Preregistered analyses showed small negative associations between wellbeing PGS and general psychopathology (β = −0.09, p<.001) and both internalizing (β = - 0.09, p<.001) and externalizing problems (β = −0.07, p<.001). ICV partially mediated the relationships between wellbeing PGS and psychopathology, accounting for 1.3-3.3% of these relationships. The findings suggest that while wellbeing PGS have limited associations with regional brain structure in children, they exhibit small protective effects against psychopathology.

**Highlights:**

- Wellbeing PGS show limited associations with brain structure in children.
- Negative association found between wellbeing PGS and parahippocampal surface area.
- Positive association found between wellbeing PGS and intracranial volume.
- Wellbeing PGS linked to lower levels of parent-reported child psychopathology.
- Intracranial volume partially mediate wellbeing PGS-psychopathology associations.

## 1. Introduction

Wellbeing is a multifaceted concept encompassing both emotional responses and cognitive judgments about life, and it plays a crucial role in overall human health and functioning (Diener et al., 2018). High levels of wellbeing have consistently been associated with better physical and mental health, as well as improved social and occupational outcomes (Bartels et al., 2013; Diener et al., 2018). However, understanding the origins and mechanisms underlying wellbeing remains challenging. While socioeconomic factors (Jebb et al., 2018), social connections and prosociality (Helliwell & Aknin, 2018), and major life events (Luhmann et al., 2012) have long been identified as key determinants of wellbeing, more recent research highlights that individual differences in wellbeing are also partly heritable and polygenic (Røysamb & Nes, 2019). Furthermore, family-based studies (Bartels, 2015; Røysamb et al., 2018; Røysamb & Nes, 2018) and genome-wide association studies (GWAS) (Baselmans, Jansen, et al., 2019; Okbay et al., 2016) show both genetic and phenotypic correlations between subjective wellbeing and related traits, such as life satisfaction, positive affect, neuroticism, and depressive symptoms, collectively referred to as the wellbeing spectrum (Baselmans, Jansen, et al., 2019).

The brain integrates a vast array of genetic and environmental influences that shape our wellbeing. Neuroscientific research can therefore provide insight into the biological basis of wellbeing. However, few studies have explored the relationship between phenotypic variation in wellbeing or closely associated concepts and brain structure. Early magnetic resonance imaging (MRI) voxel-based morphometry studies primarily found positive associations between wellbeing and regional gray matter volume (Kong et al., 2015; Lewis et al., 2014; Matsunaga et al., 2016; Sato et al., 2015; Yang et al., 2013), although some studies also found negative associations (Kong et al., 2015; Takeuchi et al., 2014). More recent findings have linked subjective wellbeing (Van ‘t Ent et al., 2017) and trait positive affect (Dennison et al., 2015) to larger hippocampal volumes, and life satisfaction to thinner lateral prefrontal cortical regions (Zhu et al., 2018). The largest study to date, involving 38,982 adult UK Biobank participants, found small significant associations between higher levels of wellbeing and greater total and regional cortical surface area, predominantly thinner regional cortex, and greater volume of several cerebellar and subcortical structures (Jamshidi et al., 2022). In contrast, extensive research on neuroticism (Bjørnebekk et al., 2013; Ferschmann et al., 2018; Holmes et al., 2012; Riccelli et al., 2017) and depression (MacSweeney et al., in press; Schmaal et al., 2016, 2017; Shen et al., 2021) has consistently found negative associations with global or regional cortical surface area, regional cortical thickness, and hippocampal volumes. Although previous studies have identified neural correlates of wellbeing, the results do not indicate whether these brain characteristics are primarily influenced by genetic factors or by environmental influences, such as experiences or lifestyle.

Polygenic scores (PGS), which capture the cumulative genetic influences from many common variants from GWAS, provide an opportunity to investigate how inherited predispositions for wellbeing relate to brain structure. Unlike phenotypic studies, this approach minimizes environmental confounding, specifically in the measurement of genetic liability. However, environmental influences may still affect brain structure and psychopathology and should be considered when interpreting associations. This approach nonetheless offers insight into potential neural mechanisms linking genetic influences on wellbeing to mental health risk and resilience. However, few studies have examined wellbeing PGS-brain structure associations. Among them, Song et al. (2019) found positive associations with cortical thickness in the right superior temporal gyrus and volume in the right insula in young adults, while Jamshidi et al. (2022) found positive associations with supramarginal cortical surface area and cerebellar volumes in middle-aged and older adults. Investigating these relationships in youth is crucial, as genetic influences on neurodevelopmental processes may shape risk or resilience for mental health problems that typically emerge in adolescence, including mood disorders, certain anxiety disorders, eating disorders, substance abuse, psychosis, and personality disorders (Blakemore, 2019; Dalsgaard et al., 2020; Whiteford et al., 2013).

To better understand the complex relationships between genetic influences on the wellbeing spectrum, brain structure, and psychopathology at the brink of adolescence, we conducted preregistered secondary analyses using data from the Adolescent Brain Cognitive Development (ABCD) study (Jernigan & Brown, 2018). We examined three primary research questions (RQ). RQ1: Is variation in wellbeing PGS associated with differences in brain structure in children? RQ2: Are there relationships between wellbeing PGS and phenotypic measures of psychopathology in children? RQ3: Does brain structure mediate the relationships between wellbeing PGS and psychopathology? Corresponding to these RQs, we formulated the following hypotheses (H) based on previous research. H1: Wellbeing PGS would be positively associated with cortical surface area and thickness in prefrontal, insular and temporal regions, as well as hippocampal volume. H2: Wellbeing PGS would be negatively associated with parent-reported general psychopathology (p-factor) and internalizing problems specifically. H3: Differences in brain structure would partly mediate the relationships between wellbeing PGS and psychopathology. Additionally, given that socioeconomic factors may moderate genetic influences on neurodevelopment and mental health (Hackman et al., 2010), we conducted exploratory analyses testing whether associations between wellbeing PGS and brain structure and psychopathology varied as a function of parental education and financial adversity. This study’s research questions, hypotheses, and statistical analyses were preregistered (https://osf.io/wrkdu), and all deviations from the preregistration are detailed in section 2.3.3.

## 2. Materials and methods

### 2.1. Participants and procedures

The current research utilized cross sectional baseline data from the Adolescent Brain Cognitive Development (ABCD) Study (https://nda.nih.gov/abcd) using the curated annual release 3.0. The ABCD Study data are stored in the NIMH Data Archive Collection #2573 and are available for approved users (Request #7474, PI: Westlye). The 3.0 release will be permanently available as a persistent data set defined in the NDA Study 901 (DOI 10.15154/1519007). The ABCD Study is an ongoing study of approximately 11,800 youths recruited at age 9-11 years in 2016-2018 across multiple sites in the United States of America, planned as a 10-year longitudinal study (Jernigan & Brown, 2018). The full protocol, including recruitment methods, exclusion criteria, and ethical approvals are described in full elsewhere (D. B. Clark et al., 2018; Garavan et al., 2018; Volkow et al., 2018). The present study was conducted in line with the Declaration of Helsinki and was approved by the Norwegian Regional Committee for Medical and Health Research Ethics (REK 2019/943).

From 11,883 available participants, we excluded 647 due to missing or poor-quality MRI data (as described below), 759 due to missing genetic data, 1 due to missing scanner serial information, and 5 duplicates. The remaining sample included 10,471 participants, including 602 monozygotic twins and 1025 dizygotic twins and siblings. To remove the issue of family relatedness, only one participant per family was retained for the analyses, yielding a final sample of 8844 participants (47.1% females) aged 8.9-11.0 years (Table 1). For the interaction analyses, 539 had missing data on parental education and 462 had missing data on financial adversity. Sensitivity analyses were as described below performed on a subsample of participants who were reported to be Non-Hispanic White.

**Table 1.** Sample descriptive statistics. N = sample size, SD = standard deviation.

| Variable | n | Mean (SD) | Min-max |
| --- | --- | --- | --- |
| Age in years | 8844 | 9.9 (0.6) | 8.9-11.0 |
| P-factor | 8842 | 6.6 (7.0) | 0.0-55.5 |
| Internalizing | 8842 | 3.8 (4.2) | 0.0-38.1 |
| Externalizing | 8842 | 4.1 (5.3) | 0.0-43.8 |
| Parental education | 8305 | 17.2 (2.5) | 3-21 |
| Financial adversity | 8382 | 0.5 (1.2) | 0-7 |

### 2.2. Measures

#### 2.2.1. Genetic data

Using summary statistics from a GWAS of wellbeing spectrum scores in adults, we estimated wellbeing PGS (Baselmans, Jansen, et al., 2019) using PRSice v2 (Choi & O’Reilly, 2019) and standard quality control procedures, including the filtering of genetic variants that are poorly imputed (INFO<.5), have high missingness (>.05), or with low minor allele frequency (MAF<.05). We based the PGS on variants passing a p=.05 significance threshold.

#### 2.2.2. Neuroimaging data

MRI data were collected across 22 sites and 29 scanners using an optimized acquisition protocol harmonized across three different types of 3T MRI scanners: Siemens Prisma, General Electric 750, and Philips. The T1-weighted image was an inversion prepared RF-spoiled gradient echo scan, using prospective motion correction when available, and with 1 mm isotropic voxel resolution. Detailed descriptions of the MRI protocol, acquisition parameters, and care and safety procedures implemented for scanning of children are presented elsewhere (Casey et al., 2018).

T1-weighted data were processed with FreeSurfer (http://surfer.nmr.mgh.harvard.edu), which performs volumetric segmentation and cortical surface reconstructions (Dale et al., 1999; Fischl et al., 1999, 2002). The image processing in the ABCD Study is described further elsewhere (Hagler et al., 2019). The images were screened for incidental findings by a neurologist, and the ABCD Study team performed manual and automated quality control (QC) of all the raw imaging data, as well as the postprocessing cortical surface reconstructions for all participants. We included the T1-weighthed data recommended by the ABCD Study team, specifically 1) no serious MRI findings, 2) T1 series passed raw QC, and 3) FreeSurfer QC not failed.

The present study used tabulated measures of cortical surface area and cortical thickness in each region of the Desikan-Killiany Atlas (2×34 variables) (Desikan et al., 2006) and volume of the following structures: accumbens, amygdala, caudate, hippocampus, pallidum, putamen, thalamus, cerebral white matter, cerebellum gray matter, and cerebellum white matter (10 variables) (Fischl et al., 2002). Measures from each hemisphere were averaged. Total intracranial volume (ICV) as estimated by FreeSurfer (Buckner et al., 2004) was used as both a covariate (in analyses on area or volume) and in exploratory analyses as an additional outcome variable.

Imaging data were harmonized across scanners prior to statistical analyses using the R package ComBat (Fortin et al., 2017, 2018; Johnson et al., 2007). This empirical Bayesian method has been adopted from genomics and applied to various brain metrics to remove unwanted sources of scan variability such as differences in MRI-scanner manufacturer while preserving biological associations in the data (Fortin et al., 2017, 2018). To ensure that biological variability was preserved during the harmonization process, the model matrix used included the following covariates: age, sex, and wellbeing PGS.

#### 2.2.3. Parent-reported child psychopathology

Continuous child psychopathology variables in the current study included general psychopathology, internalizing problems, and externalizing problems (3 variables). These measures were calculated by multiplying each scale score by its respective factor loading from a latent factor model reported on the same sample elsewhere (D. A. Clark et al., 2021), then summing the relevant weighted scores. Specifically, the model was fitted on the ABCD Study baseline parent-rated Child Behavior Checklist (CBCL) data. The CBCL (Achenbach & Ruffle, 2000) contains 119 items in which parents are presented with a particular behavior and rate the extent to which that behavior was characteristic of their child over the past 6 months on a 3-point scale. For our study, we used a higher order factor model with a priori CBCL scales as input (model number GFP-13 in (D. A. Clark et al., 2021)). In this context, “scales as input” refers to the use of the predefined CBCL syndrome scales rather than individual CBCL item scores. Higher order models posit an explicitly hierarchical structure in which observed indicators load on a number of lower order factors, which in turn load on a single higher order factor. The single general factor of psychopathology (p-factor) accounts for covariation across all symptoms, reflecting broad liability to various forms of psychopathology (Caspi et al., 2014; Caspi & Moffitt, 2018; Michelini et al., 2019). The lower order factors of internalizing and externalizing problems capture unresidualized clusters of risk for a narrower range of symptoms (D. A. Clark et al., 2021). This approach offers a robust way to capture broad and domain-specific psychopathology dimensions in children.

#### 2.2.4. Socioeconomic status

Socioeconomic status was assessed as i) highest parental education which was the highest attained education reported in number of years by at least one of the caretakers, and ii) financial adversity measured by the Parent-Reported Financial Adversity Questionnaire (PRFQ) (Diemer et al., 2013) which yields a sum score reflecting experiences of financial difficulties in the past 12 months.

### 2.3. Statistical analyses

#### 2.3.1. Main analyses

All statistical analyses were performed in R (R Core Team, 2021) Version 4.3.2, in RStudio (RStudio Team, 2020). Pearson’s correlation was used to assess associations among variables of interest. To address RQ1 and RQ2 examining the associations between wellbeing PGS and i) brain structure, and ii) measures of psychopathology, respectively, we employed a series of linear regression models.

Specifically, to address RQ1, each brain region of interest was regressed on wellbeing PGS, sex, age, 20 population components, and genotype batch. ICV was included as additional covariates in analyses where the dependent variable was cortical surface area or brain volume. Since analyses were ran with 78 outcome variables, we employed Matrix Spectral Decomposition (matSpD, https://research.qut.edu.au/sgel/software/matspd-local-version) to correct for multiple comparisons. MatSpD estimates the number of independent variables in the dataset based on the eigenvalues of the observed correlation matrix. The higher the correlation between the included measures, the lower the number of independent observations. MatSpD yields an adjusted p-value required to keep Type I Error Rate at 5%. The estimated number of independent variables was 23.057.

To address RQ2, three separate analyses were performed where psychopathology (p-factor, internalizing, externalizing) were regressed on wellbeing PGS, sex, age, 20 population components, and genotype batch. Bonferroni correction, i.e. 0.05/3 = 0.0167, was applied to correct for multiple comparisons.

To address RQ3, the R package *mediation* (Tingley et al., 2014) version 4.5.0 was used to explore whether brain structure mediated the associations between wellbeing PGS and child psychopathology. In line with the preregistration, this was only done for brain features with significant associations in RQ1. The indirect effect of wellbeing PGS on psychopathology was calculated as the product of two standardized regression coefficients extracted from a) an equation regressing psychopathology on wellbeing PGS and b) an equation regressing psychopathology on wellbeing PGS and brain structure. The same covariates were included in the equations as when testing RQ1 and RQ2. To obtain confidence intervals and test the statistical significance of the indirect effect, we applied a nonparametric bootstrapping method with 5000 iterations. We report Average Causal Mediation Estimates (ACME), and Proportion Mediated estimate, that reflects the mediation effect size. The robustness of the findings was assessed by performing a sensitivity analysis using the *medsens* function. This function measures how robust the ACME estimates are to unobserved confounders. Specifically, this analytical step identifies the sensitivity parameter rho (*ρ*) at which ACME = 0, with small *ρ* indicating that a statistically significant ACME is not robust and would not be statistically significant in the presence of a unobserved confounder causing even a small correlation between errors for the mediator and the outcome models (Chi et al., 2022).

Finally, as preregistered, exploratory analyses were performed to examine whether the strength of the associations identified in RQ1 and RQ2 varied as a function of parental education and financial adversity. In separate analyses we tested for interactions between wellbeing PGS and parental education and financial adversity on child brain structure and psychopathology, with sex, age, 20 population components, and genotype batch as covariates.

#### 2.3.2. Sensitivity analyses

As preregistered, sensitivity analyses were performed for our main RQs on a subset of participants who reported to be Non-Hispanic White (n = 4665) because of the sample compositions of the discovery GWAS and evidence that polygenic scores do not necessarily generalize across ancestries (Martin et al., 2019). As in the main analyses, MatSpD was used as a method of correction for multiple comparisons. The estimated number of independent variables in these sensitivity analyses including unrelated participants who reported to be Non-Hispanic Whites was 23.571.

#### 2.3.3. Deviations from preregistration

First, contrary to the preregistration, mixed-effects models controlling for twin and sibling status were not used to explore RQ1 and RQ2 due to model convergence issues. Instead, to remove family relatedness in the data, we only retained one participant per family and employed linear regression analyses. For each model, we extracted residuals and assessed whether the linear regression model assumptions were satisfied. When investigating RQ2, it was necessary to transform the outcome variables (internalizing, externalizing and p-factor) and log transformation yielded results that best met the linear regression model assumptions. Second, we did not include site as a random factor because literature on longitudinal modeling suggests that it is not likely that units of nesting such as site or individual scanners in multisite imaging studies such as the ABCD Study meet the theoretical assumption of a random factor (McCormick et al., 2023; McNeish et al., 2017). Instead, ComBat (Fortin et al., 2017, 2018) was used to remove non-biological variance originating from differences among MRI scanners. Third, we did not include categorical ethnicity variables as covariates since we did not have a justifiable reason to do so (Cardenas-Iniguez & Gonzalez, 2024). Finally, in addition to the preregistered analyses where ICV was included as a covariate, we extended RQ1 to also include ICV as an outcome. ICV is a heritable trait (Stein et al., 2012) which has been considered a general marker of neurodevelopment (Walhovd et al., 2022) and associated with externalizing problems in adolescents (Bashford-Largo et al., 2024).

## 3. Results

### 3.1. Descriptive statistics

Sample descriptive statistics are provided in Table 1 and density plots of study variables are shown in Figure 1. Pearson’s correlations among variables are reported in Table 2 and showed small negative associations between wellbeing PGS and psychopathology scores (range r = −.05 to −.09) and strong positive associations between the psychopathology scores (range r = .59 to .92). Moreover, wellbeing PGS was positively associated with parental education (r = .23) and negatively associated with financial adversity (r = −.14). Associations between wellbeing PGS and outcome measures of interest are plotted in Supplementary Figure 1.

**Figure 1.**
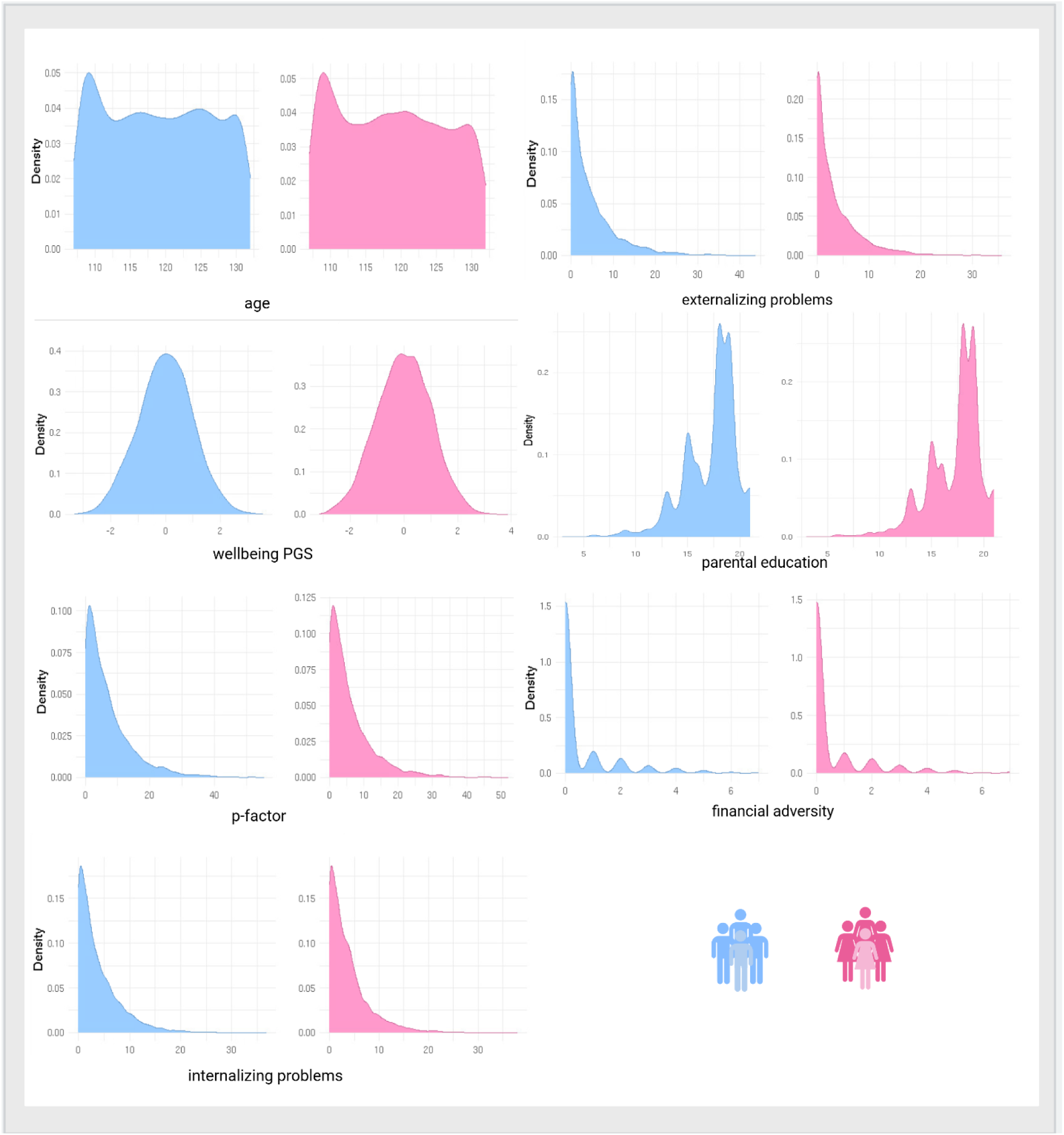
Density plots of study variables for males (blue) and females (pink)

**Table 2.** Correlations between study variables. *** *p* < .001, ** *p* < .005, *\* p* < .05.

| Variable | Age | Sex | Wellbeing<br>g PGS | P-factor | Internalizing | Externalizing | Parental<br>education<br>n | Financial<br>adversity |
| --- | --- | --- | --- | --- | --- | --- | --- | --- |
| Age | - |  |  |  |  |  |  |  |
| Sex | -.02 | - |  |  |  |  |  |  |
| Wellbeing<br>PGS | .01 | < -.01 | - |  |  |  |  |  |
| P-factor | -.02* | -.07*** | -.08*** | - |  |  |  |  |
| Internalizing | <-.01 | -.01 | -.05*** | .86*** | - |  |  |  |
| Externalizing | -.03** | -.11*** | -.09*** | .92*** | .59*** | - |  |  |
| Parental<br>education | .01 | < -.01 | .23*** | -.10*** | -.05*** | -.12*** | - |  |
| Financial<br>adversity | <.01 | < .01 | -.14*** | .20*** | .17*** | .19*** | -.30*** | - |

### 3.2. Associations between wellbeing PGS and brain structure

Controlling for age, sex, 20 population components, genetic batch, and ICV (for analyses on area and volume measures), a series of preregistered analyses showed no significant associations between wellbeing PGS and regional cortical thickness or subcortical volumes after correcting for multiple comparisons (Tables 3-4). For cortical surface area, only one region showed a corrected significant association: wellbeing PGS was negatively associated with area of the parahippocampal cortex (β = −0.03, 95% CI [−0.05, −0.01], *p* = .002). In a non-preregistered analysis, we found a corrected positive association between wellbeing PGS and ICV (β = 0.04, 95% CI [0.02, 0.06], *p* < 0.001). MatSpD computed the corrected significance threshold required to keep Type I Error Rate at 5% in this sample to be 0.0034.

**Table 3.** Beta values (unstandardized) and p-values for wellbeing PGS effects on regional cortical surface area and thickness, controlling for sex, age, 20 population components, genotype batch, and intracranial volume (for analyses on area).

| Cortical region | Surface area |  | Thickness |  |
| --- | --- | --- | --- | --- |
|  | B | p | B | p |
| Bankssts | -1.365 | .438 | 0.004 | .063 |
| Caudalanteriorcingulate | -1.623 | .192 | 0.002 | .369 |
| Caudalmiddlefrontal | 1.912 | .683 | <-0.001 | .978 |
| Cuneus | -0.182 | .933 | 0.022 | .048 |
| Entorhinal | 0.521 | .542 | -0.002 | .640 |
| Fusiform | 2.230 | .564 | 0.002 | .236 |
| Inferiorparietal | -3.964 | .609 | 0.004 | .047 |
| Inferiortemporal | 3.178 | .509 | 0.003 | .164 |
| Isthmuscingulate | 1.168 | .517 | -0.002 | .302 |
| Lateraloccipital | -0.221 | .973 | 0.002 | .308 |
| Lateralorbitofrontal | 2.791 | .334 | 0.004 | .027 |
| Lingual | -4.336 | .309 | 0.001 | .735 |
| Medialorbitofrontal | 3.275 | .129 | 0.003 | .148 |
| Middletemporal | 0.467 | .920 | 0.004 | .086 |
| Parahippocampal | -3.773 | .002 | 0.004 | .132 |
| Paracentral | -0.427 | .846 | 0.003 | .077 |
| Parsopercularis | -3.764 | .179 | 0.002 | .163 |
| Parsorbitalis | 0.729 | .421 | 0.005 | .030 |
| Parstriangularis | -0.112 | .965 | 0.003 | .111 |
| Pericalcarine | -0.088 | .973 | 0.002 | .214 |
| Postcentral | -3.995 | .482 | <0.001 | .633 |
| Posteriorcingulate | -1.041 | .591 | <0.001 | .921 |
| Precentral | 3.178 | .621 | 0.003 | .062 |
| Precuneus | -0.944 | .845 | 0.002 | .208 |
| Rostralanteriorcingulate | 0.605 | .635 | 0.002 | .483 |
| Rostralmiddlefrontal | 12.671 | .153 | 0.002 | .281 |
| Superiorfrontal | 8.202 | .358 | 0.002 | .406 |
| Superiorparietal | -0.170 | .982 | 0.002 | .214 |
| Superiortemporal | -0.355 | .941 | 0.002 | .274 |
| Supramarginal | 10.232 | .111 | 0.002 | .300 |
| Frontalpole | 0.260 | .531 | 0.003 | .359 |
| Temporalpole | 0.356 | .543 | <-0.001 | .930 |
| Transversetemporal | 0.017 | .980 | -0.001 | .525 |
| Insula | 4.686 | .036 | 0.002 | .151 |

**Table 4.** Beta values (unstandardized) and p-values for wellbeing PGS effects on brain volumes, controlling for sex, age, 20 population components, genotype batch, and intracranial volume.

| Subcortical region | B | p |
| --- | --- | --- |
| Accumbens | 1.184 | .290 |
| Amygdala | 0.438 | .836 |
| Caudate | -1.396 | .807 |
| Hippocampus | 4.397 | .267 |
| Pallidum | -1.102 | .603 |
| Putamen | -1.932 | .774 |
| Thalamus | 4.757 | .396 |
| Cerebral white matter | 111.254 | .558 |
| Cerebellum gray matter | 46.715 | .375 |
| Cerebellum white matter | -3.431 | .875 |

In a follow-up sensitivity analysis on a subset of participants who reported to be Non-Hispanic White (n = 4665), the associations between wellbeing PGS and parahippocampal surface area (β = −0.03, 95% CI [−0.05, −0.01], *p* = .017) and ICV (β = 0.04, 95% CI [0.01, 0.06], *p* = 0.006) remained of similar strength but were not significant after correction for multiple comparison. In this reduced sample, MatSpD computed the corrected significance threshold required to keep Type I Error Rate at 5% as 0.0034.

### 3.3. Associations between wellbeing PGS and child psychopathology

Controlling for age, sex, 20 population components, and genetic batch, wellbeing PGS was negatively associated with the p-factor (β = −.09, 95% CI [−0.11, −0.06], *p* < .001), internalizing problems (β = −.09, 95% CI [−0.12, −0.06], *p* < .001), and externalizing problems (β = −.07, 95% CI [−0.10, −0.05], *p* < .001). All associations were significant after Bonferroni correction for multiple comparisons. Sensitivity analyses with the subsample consisting of participants who reported to be Non-Hispanic White yielded similar results for p-factor (β = −.10, 95% CI [−0.12, −0.07], *p* < .001), internalizing (β = −.09, 95% CI [−0.11, −0.06], *p* < .001), and externalizing (β = −.08, 95% CI [−0.11, −0.05], *p* < .001).

### 3.4. Brain structure as the mediator between wellbeing PGS and psychopathology

Since wellbeing PGS in the main analyses was significantly associated only with parahippocampal surface area and ICV, mediation analyses were performed only with these two measures as potential mediators of the associations between wellbeing PGS and child psychopathology. The results of the mediation analyses showed that the indirect effects of wellbeing PGS through parahippocampal surface area on p-factor (ACME = 0.001, 95% CI [−0.002, −0.00], *p* = 0.18), internalizing problems (ACME = 0.0004, 95% CI [−0.0002, −0.00], *p* = 0.22), and externalizing problems (ACME = 0.001, 95% CI [0.002, −0.00], *p* = 0.18) were not significant. However, the mediation analyses showed that there was an indirect effect of wellbeing PGS through ICV on p-factor (ACME = −0.002, 95% CI [−0.004, 0.00], p < .001), internalizing problems (ACME = −0.001, 95% CI [−0.003, 0.00], p = 0.012), and externalizing problems (ACME = −0.002, 95% CI [−0.004, 0.00], p < .001) (Figure 2). The mediating effect (proportion mediated) of ICV explained 2.3% of the association between wellbeing PGS and p-factor, 1.3% of the relationship between wellbeing PGS and internalizing problems, and 3.3% of the relationship between wellbeing PGS and externalizing problems. ACME estimates appeared not to be robust to unobserved confounders, with sensitivity parameter rho (*ρ*) at which ACME = 0 being around −0.05 for p-factor and internalizing and externalizing problems. This means that even though there were small statistically significant mediating effects of wellbeing PGS on psychopathology though ICV, the associations were not robust and even small unmeasured confounding could explain the effect.

**Figure 2.**
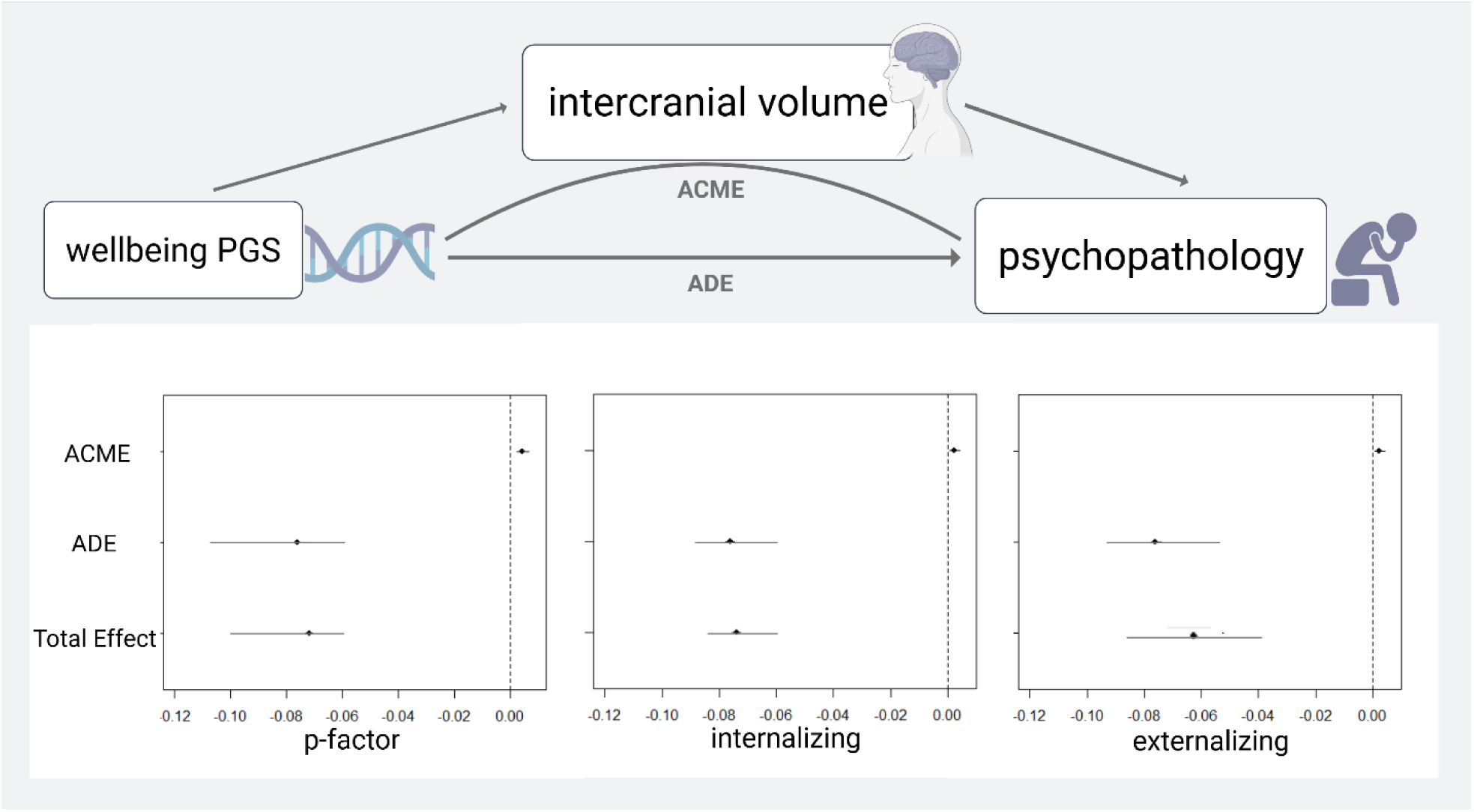
Mediation analyses exploring ICV as a mediator of the association between wellbeing PGS and child psychopathology (p-factor, internalizing problems, and externalizing problems). Significance was tested using a bootstrapping procedure with 5000 iterations. ACME = Average causal mediation effect (indirect effect). ADE = average direct effect. 95% confidence intervals are depicted for ACME, ADE, and total effect.

In follow-up sensitivity analyses on the subset of participants who reported to be Non-Hispanic White, we explored whether ICV mediated the associations between wellbeing PGS and child psychopathology. The results were similar as in the main analyses: p-factor (ACME = −0.002, 95% CI [−0.004, 0.00], p =0.009), internalizing problems (ACME = −0.001, 95% CI [−0.003, 0.00], p =0.09), and externalizing problems (ACME = −0.003, 95% CI [−0.001, 0.00], p = 0.009). The mediating effects of ICV explained 2% of the relationship between wellbeing PGS and p-factor, 1% between wellbeing PGS and internalizing problems, and 3.2% between wellbeing PGS and externalizing problems. Also here, ACME estimates appeared not to be robust to unobserved confounders, with sensitivity parameter rho (*ρ*) at which ACME = 0 being around −0.05 for p-factor and internalizing and externalizing problems.

### 3.5. Interactions with parental education and financial adversity

As preregistered, we tested whether significant associations between wellbeing PGS and outcomes of interests differed as a function of parental education and financial adversity. The interaction terms between wellbeing PGS and parental education and financial adversity on parahippocampal surface area were not significant ((β = −0.01, 95% CI [−0.03, 0.01], p = 0.222) and (β = 0.003, 95% CI [−0.02, 0.02], p = 0.728), respectively). We found significant interactions of wellbeing PGS and parental education on ICV (β = 0.03, 95% CI [0.01, 0.04], p = 0.016), p-factor (β = −0.03, 95% CI [−0.06, −0.01], p = 0.004), internalizing problems (β = −0.03, 95% CI [−0.06, −0.01], p = 0.005), and externalizing problems (β = −0.03, 95% CI [−0.05, −0.01], p = 0.019). Significant interactions of wellbeing PGS and financial adversity were only found for ICV (β = −0.02, 95% CI [−0.04, −0.001], p = 0.040). Higher parental education was associated with a stronger positive association between wellbeing PGS and ICV and a stronger negative association between wellbeing PGS and child psychopathology, while financial adversity was associated with a weaker positive association between wellbeing PGS and ICV (Figure 3).

**Figure 3.**
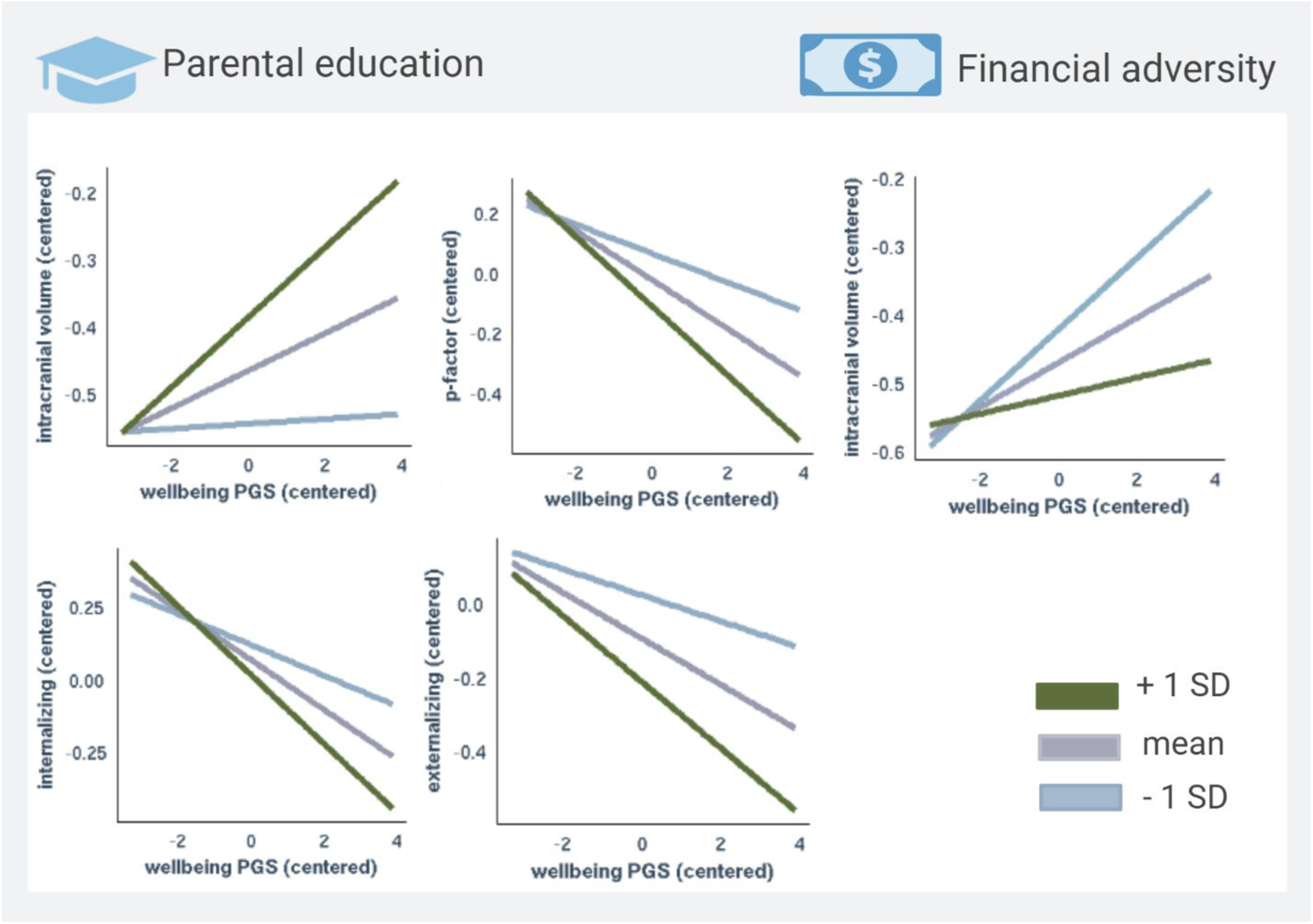
Interactions analyses between wellbeing PGS and parental education and financial adversity on ICV and child psychopathology. Linear regression analyses were performed with continuous measures of parental education and financial adversity, and with sex, age, 20 population components, and genotype batch as covariates. For illustration purposes, the sample was split into three groups based on standard deviations in parental education and financial adversity.

## 4. Discussion

This study examined the relationships between genetic predispositions for wellbeing, brain structure, and psychopathology in a large sample of children and revealed several notable findings. First, preregistered analyses showed no significant associations between wellbeing PGS and regional cortical thickness or subcortical volumes, and only a small negative association with surface area of the parahippocampal cortex. However, an exploratory analysis identified a significant positive association with ICV. Second, preregistered analyses showed that wellbeing PGS were associated with less parent-reported psychopathology in children. Third, ICV emerged as a significant mediator of the relationship between wellbeing PGS and psychopathology phenotypes. Together, these findings suggest that a genetic propensity for wellbeing provides modest protective effects against early-life psychopathology, partially mediated through general neurodevelopmental processes.

Our first hypothesis was that wellbeing PGS would be positively associated with cortical surface area and thickness in prefrontal, insular, and temporal regions, as well as hippocampal volume. This expectation was based on previous studies of brain structural correlates of phenotypic wellbeing and related concepts (De Vries et al., 2023; Jamshidi et al., 2022), neuroticism (Bjørnebekk et al., 2013; Riccelli et al., 2017), or depression (MacSweeney et al., in press; Schmaal et al., 2020), as well as a small number of genetically informed wellbeing-brain structure studies (Jamshidi et al., 2022; Song et al., 2019; Van ‘t Ent et al., 2017). These regions have been implicated in emotion regulation, stress resilience, and cognitive processes relevant to wellbeing (Ahmed et al., 2015; Berretz et al., 2021). While previous studies have shown neural correlates of phenotypic wellbeing, such studies do not distinguish genetic from environmental contributions. Our preregistered analyses, however, revealed no significant associations between wellbeing PGS and regional cortical thickness or subcortical volumes, and only a small negative association with surface area of the parahippocampal cortex, and did thus not support this hypothesis. These results contrast with prior studies in adults that have reported positive associations between wellbeing PGS and cortical thickness in the right superior temporal gyrus and volume in the right insula (Song et al., 2019) and supramarginal cortical surface area and cerebellar volumes (Jamshidi et al., 2022). Our findings do however align with studies that have found only limited associations between neuroticism or depression PGS and brain structure in healthy children (Fernandez-Cabello et al., 2022; Norbom et al., 2020; Pine et al., 2023).

In an exploratory analysis, however, we identified a small but significant positive association between wellbeing PGS and ICV. Sensitivity analyses restricted to participants who reported to be Non-Hispanic White showed similar effect sizes, although the association was only marginally significant after correction for multiple comparisons. The observed association with ICV indicates that wellbeing PGS exert a small general effect on neurodevelopment, as ICV development stabilizes in adolescence (Mills et al., 2016).

The limited associations between wellbeing PGS and brain structure in children may be explained by several factors. First, other genetic (Fernandez-Cabello et al., 2022; Grasby et al., 2020; Merz et al., 2022) and environmental (Alnæs et al., 2020; Lindseth et al., 2024; Norbom et al., 2024) factors likely play greater roles in shaping child brain structure. Second, the genetic signal captured by the wellbeing spectrum PGS (Baselmans, Jansen, et al., 2019) explains only small proportions of the heritability of the related complex traits, approximately in the 1-2% range, limiting its ability to detect associations with brain structure. Third, it is possible that associations between PGS and brain structure emerge later in life through dynamic interactions between genetic predispositions and the environment (Tucker-Drob et al., 2013).

Consistent with our second hypothesis, higher wellbeing PGS were associated with lower parent-reported psychopathology scores, also in sensitivity analyses considering Non-Hispanic White participants only. However, while we hypothesized that the association would be strongest for internalizing problems, the results showed similar effect sizes for p-factor, externalizing problems, and internalizing problems. These findings extend previous results in adults (Baselmans, Van De Weijer, et al., 2019), and suggest that wellbeing PGS confer small, non-specific negative associations with general psychopathology in children.

Our study next examined whether genetic influences on wellbeing shape neurodevelopmental processes, which in turn contribute to variation in child mental health outcomes. We found that ICV mediated the associations between wellbeing PGS and psychopathology scores, accounting for 1.3-3.3% of these relationships. This supports our third hypothesis that neurodevelopmental processes partially mediate the link between wellbeing PGS and psychopathology, albeit not through the hypothesized regional brain measures. The finding is consistent with recent studies showing that global brain volume metrics indirectly link polygenic liability to conduct problems (Lahey et al., 2024), attention-deficit hyperactivity disorder (Mooney et al., 2021), and psychotic-like experiences (Karcher et al., 2022). Our current cross-sectional findings should however be interpreted with caution. First, the estimates were not robust and thus warrant replication. Second, the causal pathways remain to be characterized in longitudinal analyses, and neurodevelopmental and disease processes may exert bidirectional effects. Finally, exploratory analyses revealed that parental education level and financial adversity moderated the associations between wellbeing PGS and key outcomes. Specifically, higher parental education was linked to a stronger positive association between wellbeing PGS and ICV, as well as stronger negative associations between wellbeing PGS and child psychopathology, while financial adversity was associated with a weaker positive association between wellbeing PGS and ICV. These findings align with prior research showing that socioeconomic status can moderate the heritability or genetic contribution on various phenotypes, most consistently cognitive abilities (Gottschling et al., 2019; Rowe et al., 1999; Tucker-Drob et al., 2011; Tucker-Drob & Bates, 2016). While the specific mechanisms remain unclear, one theory is that enriched environments that have fewer external constraints, which are often associated with higher SES (e.g., higher parental education), provide more opportunities for genetic potential to manifest through transactional processes (Tucker-Drob et al., 2013). Conversely, the weaker association between wellbeing PGS and ICV in context of financial adversity may reflect a more constrained developmental environment, where limited access to enriching experiences and heightened environmental stressors reduce the expression of genetic predispositions. However, these findings should be interpreted with caution, as the relatively limited variance in some socioeconomic measures, particularly financial adversity, within the ABCD Study may limit generalizability to more socioeconomically diverse populations.

This study has several limitations. First, the absence of phenotypic wellbeing measures in the sample precluded an evaluation of whether the wellbeing PGS predicts subjective wellbeing in children. Given the challenges of measuring wellbeing in this age group (Aked & Thompson, 2009), future studies should incorporate developmentally appropriate assessments to clarify this link. Second, although sensitivity analyses were conducted in Non-Hispanic White participants, the wellbeing PGS was derived primarily from European ancestry GWAS data, limiting its predictive power in diverse populations. Expanding ancestry-inclusive genetic studies is essential for improving generalizability. Third, the cross-sectional design precludes conclusions about developmental timing. Longitudinal research is needed to examine how genetic influences on brain structure and psychopathology unfold across development.

In summary, this study provides novel insights into the relationships between genetic predispositions for wellbeing, brain structure, and childhood psychopathology. While preregistered analyses showed limited associations between wellbeing PGS and regional brain structure, exploratory findings suggest a modest link to global neurodevelopmental processes, as reflected in ICV. Further, the protective effects of wellbeing PGS against psychopathology appear partially mediated by ICV and are moderated by environmental factors such as parental education and financial adversity.

## CRediT authorship contribution statement

**Christian K. Tamnes:** Writing – original draft, Writing – review & editing, Conceptualization, Funding acquisition. **Andreas Dahl:** Writing – review & editing, Formal analysis. **Dennis van der Meer:** Writing – review & editing, Formal analysis. **Ingrid Agartz:** Writing –review & editing. **Dag Alnaes:** Writing – review & editing. **Ole A. Andreassen:** Writing – review & editing. **Kathryn L. Mills:** Writing – review & editing. **Linn B. Norbom:** Writing – review & editing. **Geneviève Richard:** Writing – review & editing. **Ronald E. Dahl:** Writing – review & editing, Funding acquisition. **Espen Røysamb:** Writing – review & editing. **Lars T. Westlye:** Writing – review & editing. **Lia Ferschmann:** Writing – original draft, Writing – review & editing, Formal analysis, Visualization.

## Declaration of Generative AI and AI-assisted technologies in the writing process

During the preparation of this work the authors used ChatGPT 4o to improve readability and language. After using this tool, the authors reviewed and edited the content as needed and takes full responsibility for the content of the publication.

## Acknowledgments

Data used in the preparation of this article were obtained from the Adolescent Brain Cognitive Development^SM^ (ABCD) Study (https://abcdstudy.org), held in the NIMH Data Archive (NDA). This is a multisite, longitudinal study designed to recruit more than 10,000 children age 9-10 and follow them over 10 years into early adulthood. The ABCD Study® is supported by the National Institutes of Health and additional federal partners under award numbers U01DA041048, U01DA050989, U01DA051016, U01DA041022, U01DA051018, U01DA051037, U01DA050987, U01DA041174, U01DA041106, U01DA041117, U01DA041028, U01DA041134, U01DA050988, U01DA051039, U01DA041156, U01DA041025, U01DA041120, U01DA051038, U01DA041148, U01DA041093, U01DA041089, U24DA041123, U24DA041147. A full list of supporters is available at https://abcdstudy.org/federal-partners.html. A listing of participating sites and a complete listing of the study investigators can be found at https://abcdstudy.org/consortium_members/. ABCD consortium investigators designed and implemented the study and/or provided data but did not necessarily participate in the analysis or writing of this report. This manuscript reflects the views of the authors and may not reflect the opinions or views of the NIH or ABCD consortium investigators.

## Data availability

Qualified researchers can request access to ABCD shared data from the NIMH Data Archive (NDA).

## Supplementary material

**Supplementary Figure 1.**
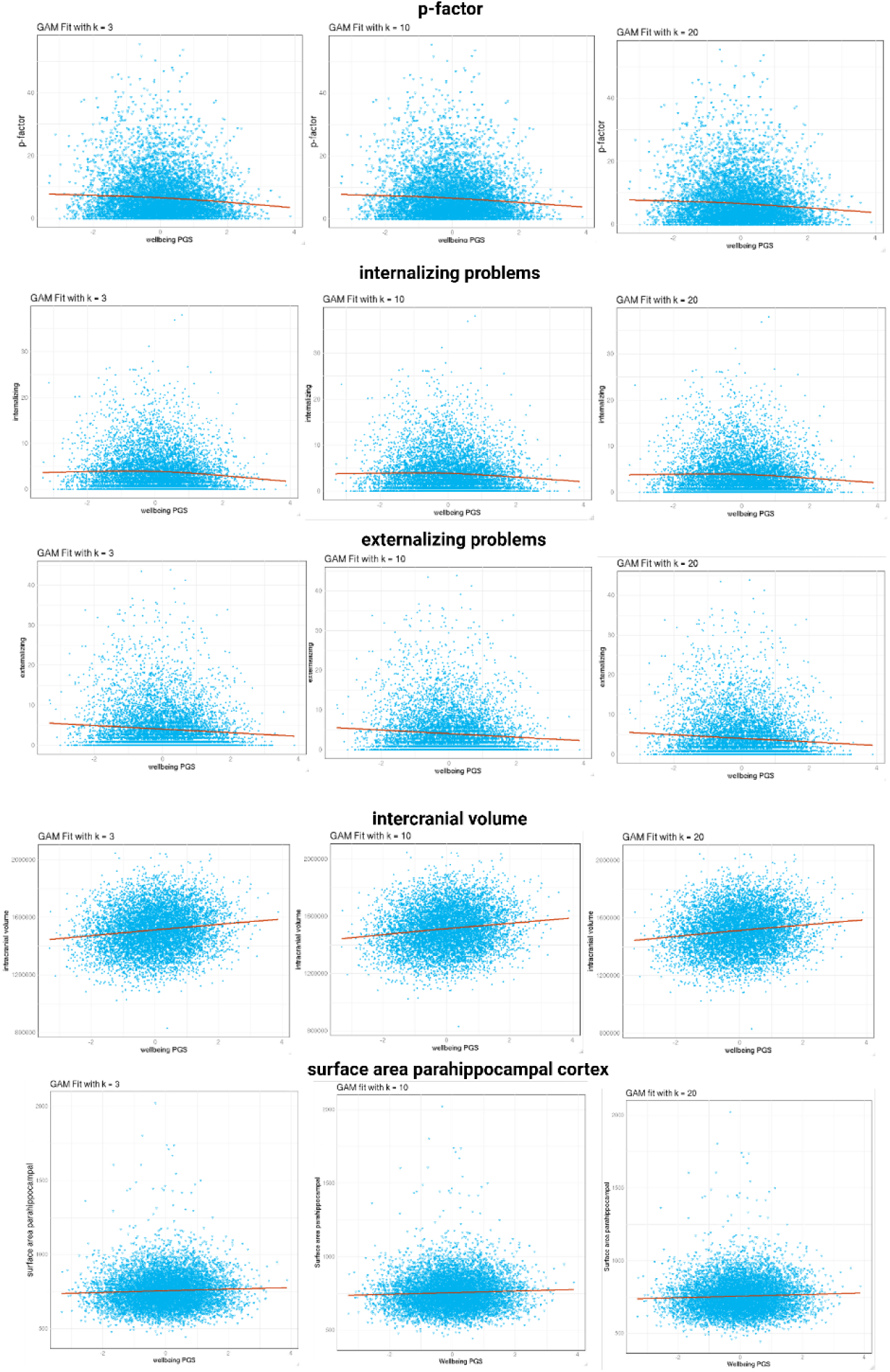
Shows associations between wellbeing PGS and 1) p-factor, 2) internalizing problems, 3) externalizing problems, 4) intracranial volume, and 5) parahippocampal surface area. Generalized additive models (Hastie & Tibshirani, 1986) with thin-plate regression splines (Wood, 2003) were applied with the mgcv package in R with different k-values (3,10, and 20) to allow for different degrees of wiggliness of the curve. Regardless of the number of splines applied, the associations appear to be largely linear.

